# Innovative Approaches to Improve Knowledge of Zoonoses among Wildlife Hunters and Traders in Epe, Lagos, Nigeria: A Community Action Network-Based Intervention

**DOI:** 10.1101/2024.02.27.24303439

**Authors:** E Cadmus, E.J. Awosanya, H.K. Adesokan, V.O. Akinseye, F. Olaleye, O Morenikeji, E.O. Fawole, Rashid Ansumana, K.O. Ayinmode, D.O. Oluwayelu, S. Cadmus

**Affiliations:** Department of Community Medicine, College of Medicine, University of Ibadan, Ibadan, Nigeria; Department of Veterinary Public Health and Preventive Medicine, University of Ibadan, Ibadan, Nigeria; Department of Chemical Sciences, Augustine University, Ilara-Epe, Lagos, Nigeria; Damien Foundation Genomic and Mycobacteria Research & Training Centre, University of Ibadan, Ibadan, Nigeria; Department of Zoology, University of Ibadan, Ibadan, Nigeria; School of Community Health Sciences, Njala University, Bo, Sierra Leone; Department of Veterinary Parasitology and Entomology, University of Ibadan, Ibadan, Nigeria; Department of Microbiology, University of Ibadan, Ibadan, Nigeria; Centre for Control and Prevention of Zoonoses, University of Ibadan, Ibadan, Nigeria; Nigeria Institute of Medical Research, Lagos, Nigeria

## Abstract

The West Africa One Health project is a multi-country project designed to utilise the One Health approach and deploy the Community Action Networks (CAN) to improve knowledge of high-risk communities on zoonoses. Majority of emerging zoonoses occur at the human-wildlife interface, of which wildlife hunters and traders are critical stakeholders. We assessed the effectiveness of a CAN-based intervention involving the use of a video documentary and case studies as model tools in improving the knowledge of zoonoses among wildlife hunters and traders in Epe, an established hunting community in Lagos State, Nigeria.

A quasi-experimental study design involving a total of 39 consenting registered wildlife stakeholders was adopted. A pre-tested, semi-structured, interviewer-administered questionnaire was used to obtain data on the participant’s sociodemographic characteristics, awareness level, and knowledge of zoonoses pre and post CAN-based intervention. Data were analysed using descriptive statistics McNemar and Wilcoxon Signed Ranks tests at a 5% level of significance.

The mean age of the participants was 46.7 ± 10.9 years. Most (76.9%) identified as male and had at least secondary education (89.7%). The number of participants who were aware that diseases could be contracted from animals and that it could be through inhalation and close contact increased significantly from 13 (33.3%), 2 (5.1%), and 9 (23.1%) pre-intervention to 37 (94.9 %), 11 (28.2%), and 21 (53.8%) post-intervention, respectively. The overall median knowledge score increased significantly from 1 (Interquartile range (IQR): 0 – 2) pre-intervention to 3 (IQR: 2 – 4) post-intervention.

The CAN-based intervention involving the use of a video documentary and case studies as model tools was effective in improving the knowledge of zoonoses among wildlife hunters and traders in the hunting community and may be beneficial for future practice.

## Introduction

Hunting of wildlife by humans is an ancient practice associated with a considerable risk for cross-species infection transmission. Bush meat processing-related activities, including hunting, trading and consumption of wildlife, prevalent in sub-Saharan Africa, have historically and currently represent a source of zoonotic disease transmission, and their potential to trigger future outbreaks remains a concern [1]. Over 75% of emerging disease pathogens are zoonotic, 60% of which spread from domestic or wild animals to humans, while 80% are of concern regarding bioterrorism [2]. Worldwide, these emerging zoonoses account for 2.5 billion cases and 2.7 billion deaths each year [2, 3, 4].

Bushmeat trade and consumption remain an important interface for the transmission of zoonotic pathogens [5, 6]. At such an interface, transmission could occur directly through skin-to-skin contact, scratches, animal bites, or contact with body fluids. Zoonosis transmission could also occur via the oral route with accidental ingestion of pathogens present in contaminated water, food and unhygienic practices [6]. The different points along the bushmeat value chain from the time of harvest, exsanguination, dehairing, de-feathering, evisceration and consumption are critical sources of direct contamination [7, 8].

In sub-Saharan Africa, in particular, there are no regulated food safety standards along the often informal trade chains from the harvesting, transportation and butchering of the animals to the preservation of meat. This lapse further exposes wildlife hunters, butchers, vendors and consumers to the transmission of zoonotic diseases and food-borne illnesses [9]. In the tropical forests of Africa, people in the local settings depend heavily on bushmeat as a cultural, economic and nutritional component of their livelihood [10]. For instance, in Nigeria, the bushmeat trade is growing at an alarming rate, with an estimated volume of 900,000 kilograms sold annually [11]. Large profit margins create incentives for the bushmeat trade across all levels of the commodity and supply chain biased towards larger and rare species, allowing bushmeat to reach national and international markets [11, 12].

Raised awareness is the most important strategy for preventing occupational diseases, yet available data show poor awareness of zoonoses among occupationally exposed individuals in Nigeria [13]. Worse still, this poor awareness is often enshrined in cultural backgrounds established over the years of practice as hunters or wild animal traders. Therefore, an intervention targeted towards achieving improved awareness and ultimately safeguarding the health of the bushmeat hunters, consumers and the general public is imperative.

The West Africa One Health (WAOH) project is a multi-country project designed to utilise the One Health approach and deployment of the Community Action Networks (CAN) to improve knowledge of high-risk communities on zoonoses. The CAN is a concept rooted in the principles of Community-Based Participatory Research (CBPR), a collaborative approach that empowers communities to engage in research from project inception [14]. The CBPR has shown success in addressing various public health issues, fostering community trust, improving data quality, and enhancing the relevance of health interventions. Its primary advantage lies in empowering communities to identify and solve their problems, aiding decision-makers and service providers in resource mobilisation and policy improvement [15, 16].

The CAN plays a critical role in community involvement in disease surveillance and has resulted in community empowerment, enhanced data quality, and more effective response to health threats [17]. The process often starts with a community needs assessment, and assessment of baseline knowledge with regards to zoonotic diseases. Information obtained may then be used for the design of targeted interventions, communication needs and subsequent knowledge transfer. Over the past years, different methods have been deployed to improve awareness of communities on zoonoses. However, the use of video documentaries and case studies has not been explored, especially within the CAN set-up.

Meanwhile, health interventions and health education via videos have been shown to provide a convenient, accessible and cost-effective method to encourage positive change and improvement in patient behaviour [18, 19]. Besides, the case study approach is widely recognised as offering an invaluable resource for understanding the dynamic and evolving influence of context on complex, system-level interventions [20–21]. The present study employed a CAN-based intervention involving the use of a video documentary and case studies as model tools for improving knowledge of zoonoses among wildlife hunters and traders in Epe, an established hunting community in Lagos State, Nigeria.

## MATERIALS AND METHODS

### Study Settings

The study area is Epe with geo-coordinates of 6°35’N and 3° 59’E (Figure 1). Epe is one of the five divisions in Lagos State, South West Nigeria. It lies within the lowland rainforest zone and is bounded by the north bank of the coastal Lagos Lagoon. The human population in Epe LGA was estimated at 269,000 in 2022 [22] and mostly of the Yoruba speaking tribe. The primary occupation is fishing; however, agriculture is also practised. Historically, one of the settlers of the town was a hunter and hunting is still commonly practised in the area. Epe is home to five higher institutions of learning and 20 Primary Health Care Centres [23]. It has both fresh, seafood and wildlife markets.

**Fig 1:**
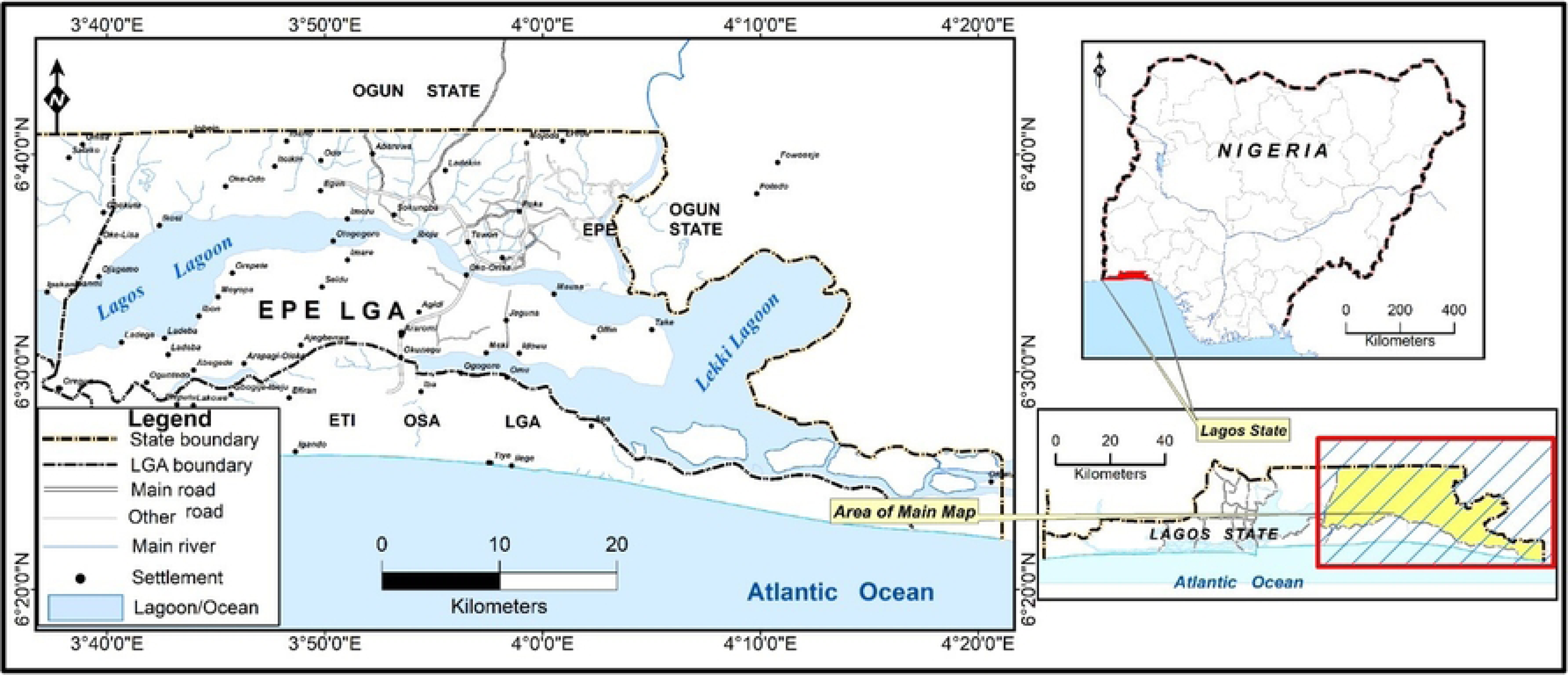
**Map of Epe Local Government areas (LGA) with Lagos State and Nigeria inset**

### Community Entry

Prior to the training, an initial visit was made to interact with the community gatekeepers, including the market leader, head hunter and youth leader. During the visit, the project aim and the modalities were explained. The process was facilitated by a member of the team, OAM, who had a pre-existing linkage with the community. The head of the hunters association assisted with the recruitment of members who were registered on their platform, while the market and youth leaders recruited the wildlife traders.

### Study Design and Period

The study adopted a quasi-experimental design and was conducted between 2^nd^ and 3^rd^ of August 2023.

### Study Participants

The study participants were wildlife hunters and traders who also doubled as processors. Members (72.5%) of the hunters association (N=40) and consenting members (34.5%) of the wildlife traders (N = 29) of the registered population in Epe town were included in the study.

### Sampling and Data Collection

Total sampling was adopted because of the finite population of hunters and wildlife traders in the study area. A total of 39 individuals, 29 hunters and 10 wildlife traders, respectively, were available and consented to participate in the study. Four Focus Group Discussions (FGD) were conducted. Each FGD consisted of nine to eleven participants and lasted between 45 to 60 minutes. The discussions were used to elicit information on precautions taken when trading or handling wildlife and self-dead animals due to idiopathic causes. Other information included the participant’s experience and perceived role in zoonotic disease surveillance and reporting, as well as prevention of the spread of zoonotic diseases. In addition, access to and promotion of healthcare services and participant views on the prioritisation of economic gain over personal health among the study participants were also evaluated.

A pre-tested, semi-structured, interviewer-administered questionnaire was also used to obtain data on the participant’s sociodemographic characteristics, awareness level, knowledge, health and safety measures among the respondents on some zoonoses of public health concerns pre-and post-intervention. The interventions included a viewing of a short documentary and discussions using case studies on zoonoses at the human-wildlife interface. The short video documentary was produced in the local language (Yoruba) and titled “IFURA”, meaning “Beware”, and had four sections.

**Section 1:** Introduction to Zoonotic Diseases: which provided an overview of wildlife and zoonotic diseases, how they spread from animals to humans, and the potential impact on public health

**Section 2:** Recognising Zoonotic Disease Symptoms: common symptoms of zoonotic diseases were highlighted, and the importance of timely medical attention was emphasised.

**Section 3:** Preventive Measures for Hunters and Bushmeat Sellers: practical preventive measures that hunters and bushmeat sellers could adopt in their daily practices, which included proper hygiene, the use of personal protective equipment (PPE), and safe animal handling techniques, were highlighted

**Section 4:** Collaboration with Local Health Authorities: the focus was on the importance of working closely with local health authorities to report suspected zoonotic cases and facilitate disease surveillance.

The documentary, which lasted 9:01 minutes, was played three times. After the first viewing of the video, there was an opportunity for participant feedback, questions and clarifications. After that, the process was repeated two more times. The session was followed by a discussion using hypothetical case studies on Mpox and anthrax, which focused on the risk of exposure, safety precautions for handling and processing wildlife, health-seeking behaviour and information dissemination.

### Ethical Consideration

Ethical approval was obtained from the University of Ibadan/University College Hospital Institutional Review Board (UI/UCH/22/0305). Informed verbal consent was obtained from the study participants, and confidentiality of the data was maintained

## Data Analysis

Data obtained from the study were coded, entered into Microsoft Excel 16.0, and analysed using SPSS version 16^®^. Thematic analysis was done for the qualitative data. Descriptive analyses such as frequencies, means, and proportions were also performed. The significant differences in the proportion of knowledge of zoonoses pre- and post-intervention among the participants were done using the McNemar test. Each item in the knowledge scale was given an equal weight of one for each correct option with a total obtainable score of 10. The significant difference in the median knowledge scores pre- and post-intervention was assessed using the Wilcoxon Signed Ranks test. The level of significance was 5%.

## RESULTS

### Sociodemographic Characteristics of Study Participants

The mean age of the study participants was 46.7 ± 10.9 years. Most (76.9%) identified as male, while more than half (56.4%) were Christians. The majority (89.7%) of the participants had secondary education. Furthermore, less than half (41.0%) of the participants reported wildlife hunting activity as a full-time occupation. A majority (87.2%) of the participants had been involved in the business of hunting or trading wildlife for over 10 years (Table 1).

**Table 1.** Sociodemographic characteristics of wildlife hunters and traders in Epe, Lagos State, Nigeria (N = 39)

|  | Frequency (n) | Percentage (%) |
| --- | --- | --- |
| **Gender** |  |  |
| Female | 9 | 23.1 |
| Male | 30 | 76.9 |
| **Age(in years)** |  |  |
| <40 | 8 | 20.5 |
| 40 – 49 | 16 | 41.0 |
| >50 | 15 | 38.5 |
| **Religion** |  |  |
| Christianity | 22 | 56.4 |
| Islam | 17 | 43.6 |
| **Years in business** |  |  |
| 1 – 5 | 2 | 5.1 |
| 6 – 10 | 3 | 7.7 |
| >10 | 34 | 87.2 |
| **Time Involved** |  |  |
| Full Time | 16 | 41.0 |
| Part-Time | 23 | 59.0 |
| **Level of education** |  |  |
| No formal | 1 | 2.6 |
| Primary | 14 | 35.9 |
| Secondary | 20 | 51.3 |
| Tertiary | 4 | 10.3 |
| **Reasons for hunting/wildlife trading** |  |  |
| Cultural/ Medical Purpose | 20 | 51.3 |
| Source of Income | 26 | 66.7 |
| Source of Protein | 20 | 51.3 |

### Knowledge of Participants on Zoonoses

About a third, 13 (33.3%), of the participants were not aware that diseases could be contracted from animals pre-intervention. However, awareness of zoonotic disease transmission increased significantly (p < 0.001) post-intervention. On the possible route of infection, only 2 (5.1%), 9 (23.1%), and 5 (12.8%) participants indicated that it could be through inhalation, close contact and eating products from infected animals, respectively, pre-intervention. The responses increased significantly post-intervention to 11 (28.2%) for inhalation (p = 0.012) and 21 (53.8%) for close contact (p = 0.004). However, there was no different significant increase in the response to eating products from infected animals 6 (15.4%). There was no significant difference in the response to symptoms of diseases in animals that they knew pre and post-intervention. Majority of the participants indicated that it is not safe to sell or eat animals that are dead from unknown causes pre- (37; 94.9%) and post- (38; 97.4%) interventions. In addition, the majority (31; 79.5%) of the respondents knew that it is unsafe to kill an animal displaying evident signs of sickness pre-intervention. However, the percentage improved significantly (37; 94.7%) following the intervention (p = 0.031) (Table 2).

**Table 2.**
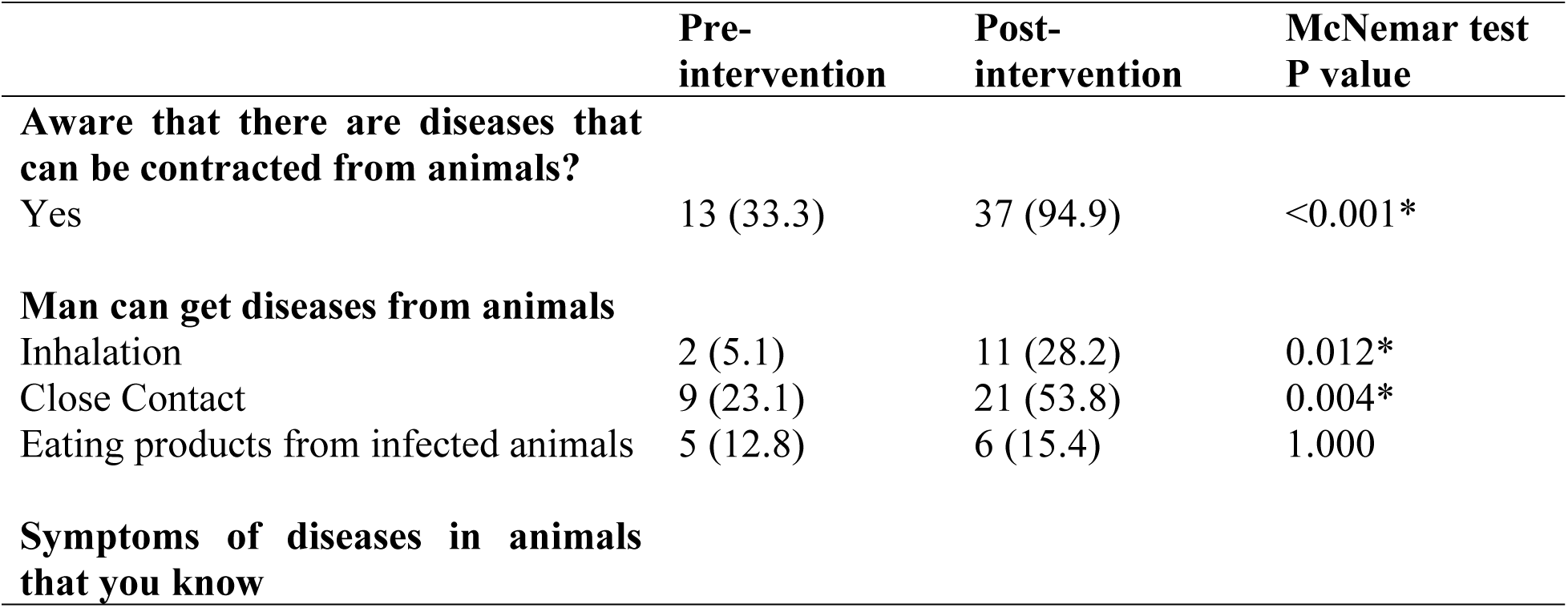
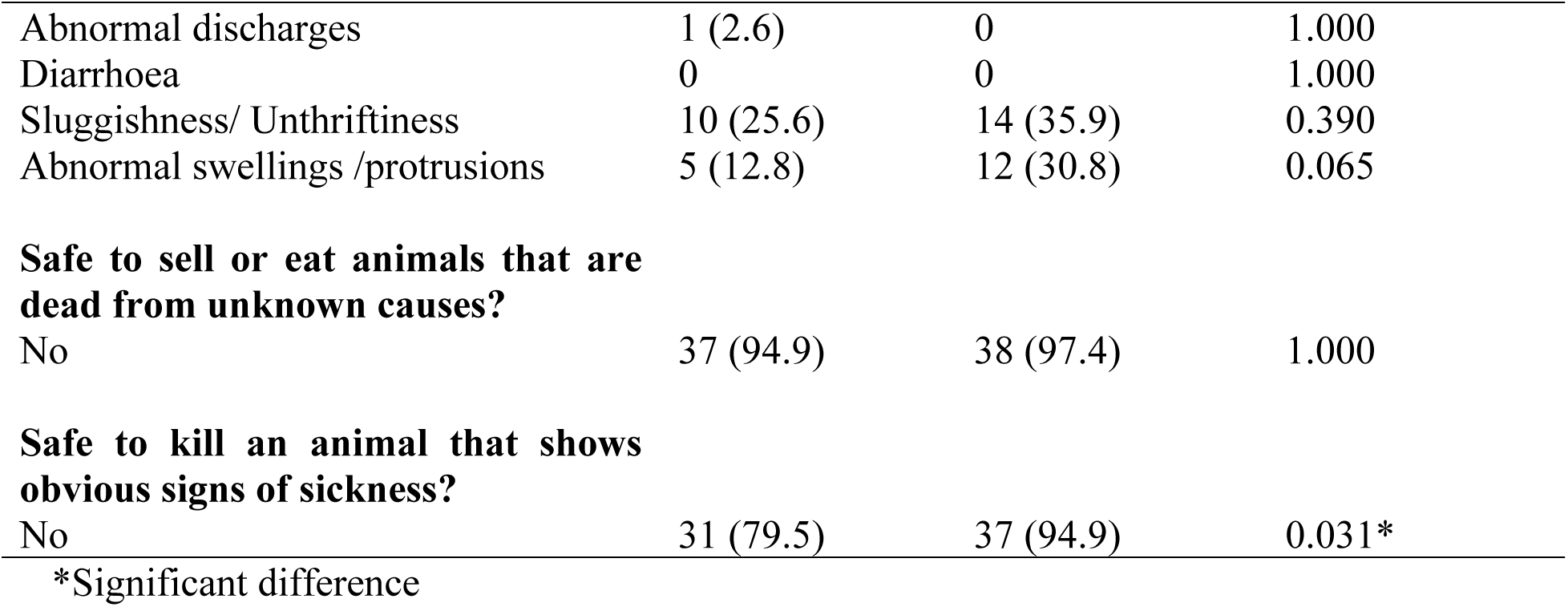
Knowledge of zoonoses pre- and post-intervention among wildlife hunters and traders in Epe, Lagos State, Nigeria (n = 39)

|  | Pre-intervention | Post-intervention | McNemar test P value |
| --- | --- | --- | --- |
| **Aware that there are diseases that can be contracted from animals?** |  |  |  |
| Yes | 13 (33.3) | 37 (94.9) | <0.001* |
| **Man can get diseases from animals** |  |  |  |
| Inhalation | 2 (5.1) | 11 (28.2) | 0.012* |
| Close Contact | 9 (23.1) | 21 (53.8) | 0.004* |
| Eating products from infected animals | 5 (12.8) | 6 (15.4) | 1.000 |
| **Symptoms of diseases in animals that you know** |  |  |  |
| Abnormal discharges | 1 (2.6) | 0 | 1.000 |
| Diarrhoea | 0 | 0 | 1.000 |
| Sluggishness/ Unthriftiness | 10 (25.6) | 14 (35.9) | 0.390 |
| Abnormal swellings /protrusions | 5 (12.8) | 12 (30.8) | 0.065 |
| **Safe to sell or eat animals that are dead from unknown causes?** |  |  |  |
| No | 37 (94.9) | 38 (97.4) | 1.000 |
| **Safe to kill an animal that shows obvious signs of sickness?** |  |  |  |
| No | 31 (79.5) | 37 (94.9) | 0.031* |

\*Significant difference

### Difference in Knowledge Before and After the Intervention

The median knowledge score, pre-intervention, was 1.00 (Interquartile range: 2). This, however, increased to 3 (Interquartile range: 2) post-intervention. The difference in the median knowledge scores pre- and post-intervention was significant (p < 0.001) (Table 3; Fig. 2).

**Fig. 2.**
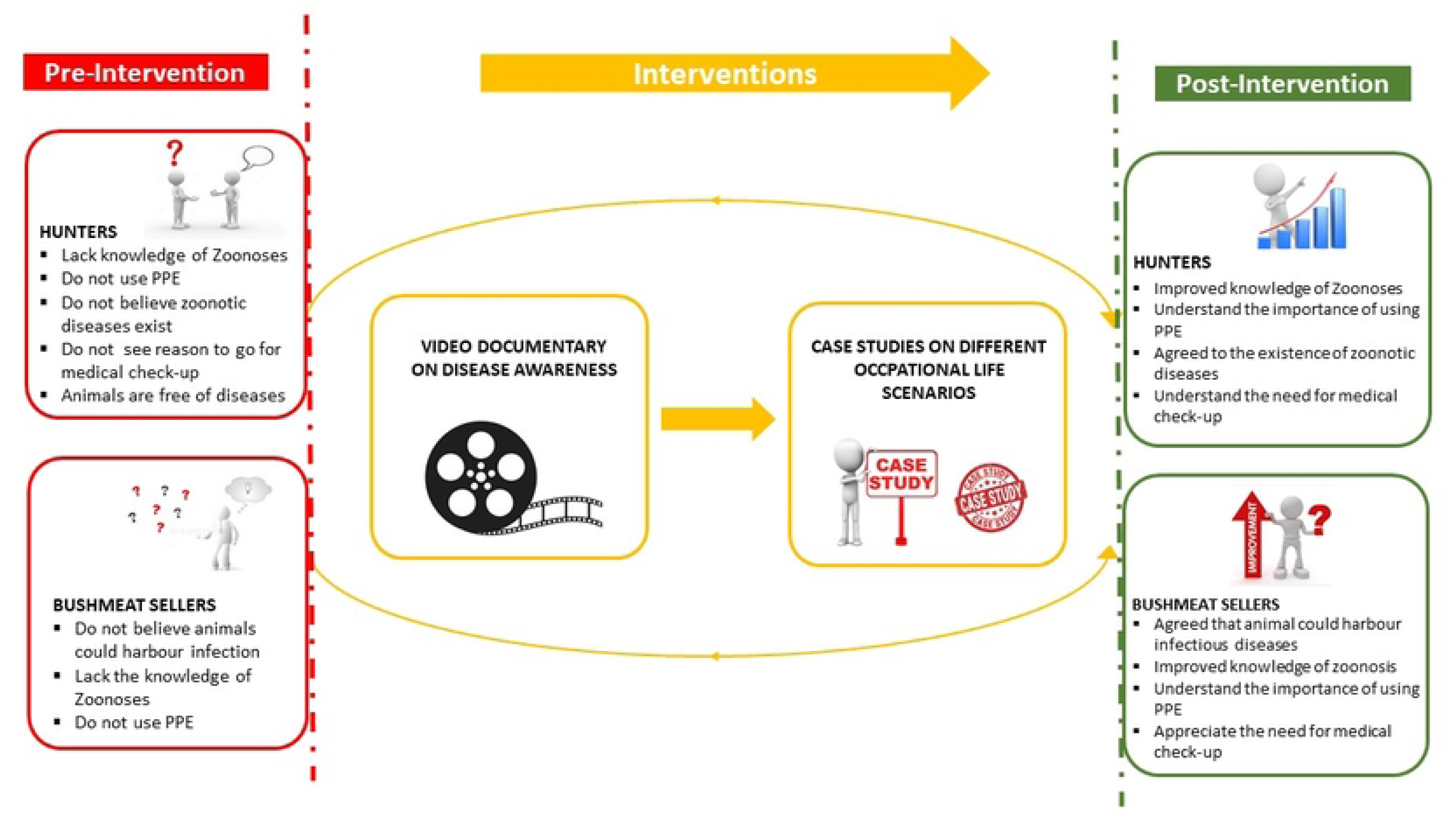
**Procedures for data collection and interventions used**

**Table 3.**
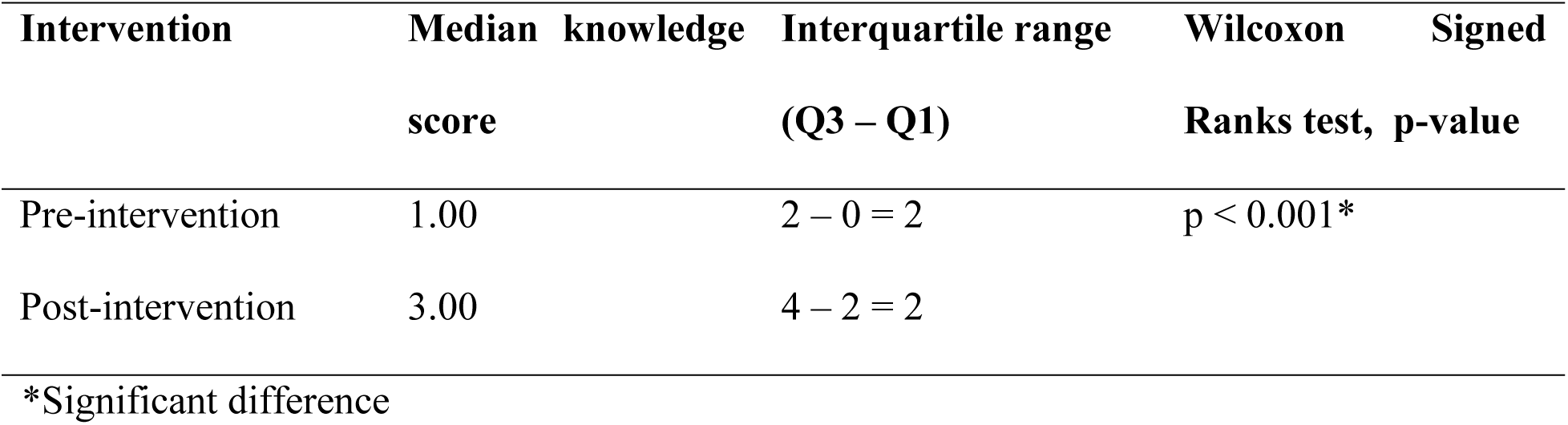
Difference in Knowledge Score pre- and post-intervention among wildlife hunters and traders in Epe, Lagos State, Nigeria (n = 39)

### Health and Safety Measures among Wildlife Hunters and Traders

All the hunters and wildlife traders had at least once or twice contact with wildlife animals, and the majority, 30 (76.9%), washed their hands immediately after such contact. The majority, 30 (76.9%), have not received any training on zoonotic diseases and their prevention. Almost half, 19 (48.7%) of the participants, have occasionally or never gone for medical check-ups or health screenings. The majority, 32 (82.1%), reported not to have experienced any work-related health issues. Although more than half (56.4%) of the participants reported a cut or injury related to their work, only 2 (9.1%) reported becoming sick after such a cut or injury. About two in every three of the participants, 24 (61.5%), reported to have seen ticks on either their body or hunting dogs. Nearly all 38 (97.4%) eat what they hunt or trade in. About all 38 (97.4%) indicated an interest in receiving training on zoonotic diseases and safe wildlife trading practices (Table 4).

**Table 4.** Health and safety measures among wildlife hunters and traders in Epe, Lagos State, Nigeria (N= 39)

|  | Frequency (n) | Percentage (%) |
| --- | --- | --- |
| **Frequency of contact with wildlife animals** |  |  |
| Always (Everyday) | 16 | 41.0 |
| Sometimes (Once/twice a week) | 18 | 46.2 |
| Often (> twice a week) | 5 | 12.8 |
| **Frequency of handwashing following animal contact** |  |  |
| Immediately | 30 | 76.9 |
| Once in a while | 8 | 20.5 |
| Never | 1 | 2.6 |
| **Received training on zoonotic diseases and prevention** |  |  |
| No | 30 | 76.9 |
| Yes | 9 | 23.1 |
| **Frequency of medical check-ups or health screenings** |  |  |
| Not at all | 14 | 35.9 |
| Occasionally | 5 | 12.8 |
| Regularly | 15 | 38.5 |
| Sometimes | 5 | 12.8 |
| **Ever experience of work-related health health issues** |  |  |
| No | 32 | 82.1 |
| Yes | 7 | 17.9 |
| **Reported work-related cuts or injuries** |  |  |
| No | 17 | 43.6 |
| Yes | 22 | 56.4 |
| **Reported sickness after work-related cut or injury (n = 22)** |  |  |
| No | 20 | 90.9 |
| Yes | 2 | 9.1 |
| **Have you ever seen ticks on yourself or animals you hunt or trade?** |  |  |
| No | 15 | 38.5 |
| Yes | 24 | 61.5 |
| **Interest in receiving training on zoonotic diseases and safe wildlife trading practices** |  |  |
| No | 1 | 2.6 |
| Yes | 38 | 97.4 |
| **Do you or your family eat what you hunt or trade?** |  |  |
| No | 1 | 2.6 |
| Yes | 38 | 97.4 |

### Qualitative Findings

The majority of participants did not believe that humans can contract diseases from wildlife. However, part of the precautions taken during hunting expeditions included the use of gloves, boots, nose masks, protective clothing, glasses, herbs, and charms, while the traders hardly took any precautions. One of the popular views among the discussants:

*“But that a wild animal would have a zoonotic disease that will affect humans, I haven’t experienced it before. Contrary to what they’re saying around in the news, it is not real. It’s not real to me.” **(Hunter, FGD 1)***

### Experience and Handling of Animals found Dead

Most of the participants have experienced the situation of self-dead animals during hunting expeditions. However, the majority would take such animals home and process them for eating, provided they could attribute the cause of death to trap or gunshot wounds, old age or killed by another animal. In situations where the participants could not ascribe the cause of death to any of the earlier reasons mentioned, if the carcass is fresh, it could as well be eaten.

As reflected by a participant*: “I have eaten dead animals that I didn’t know how they died before. It is even usually very delicious. If I see a dead animal that is fresh and not rotten, it is suitable for eating for me.”**(Hunter FGD 2)***

Exceptions mentioned included when putrefaction has set in the carcasses or suspected death due to snake bite. On the disposal of a carcass not suitable for consumption, few would bury it, but the majority would leave it to decompose in the open.

### Participants’ Perceived Roles in Zoonotic Disease Surveillance and Reporting and Contribution to Public Health

The discussants did not perceive they had a role to play in disease reporting because they were not convinced that diseases were transmissible between humans and wildlife. As a participant emphasised during the discussion:

*“We don’t even experience the spread of diseases, so we cannot report what we do not have.**” (Hunter FGD1)***

However, they indicated their role in upholding communal security and reporting any threat to the community due to the incursion of dangerous animals such as pythons, lions and hyenas. Also, most agreed that they have roles to play in promoting public health, which include proper disposal of inedible offal through burying or service of waste management truck in order to prevent environmental pollution and disease spread by flies; proper processing and cooking of wildlife; non-sales of putrefying meat.

### Awareness and Personal Experience on Zoonoses and Access to Promotion of Healthcare Services

The unanimous position was that all have neither witnessed nor heard of any incident where zoonotic diseases were transmitted within their trade or to consumers of their products. The general view was that the government should improve the healthcare delivery services to their group. As such, the healthcare system would be better positioned to support hunters and wildlife traders, not in terms of zoonotic disease prevention, but access to healthcare.

In one of their views:

“Although hunters take herbs often, modern medicine is also important.”

*“I would like the government to provide good healthcare for those that sell bush meat, teaching them about proper hygiene.” **(Trader, FGD4)***

### View on Prioritisation of Economic Gain over Personal Health

All agreed that health is more important than economic gain, as expressed by one of the discussants, “*if you are not healthy, you cannot work*.” ***(Hunter FGD2)***

## Discussion

Improving knowledge, attitudes, and behaviours regarding zoonotic diseases among high-risk groups, such as wildlife hunters and traders, is vital for the formulation and effective implementation of appropriate disease prevention and control strategies in any given area [24]. The CAN-based intervention using model tools of video documentaries and case studies employed in the present study significantly enhanced the knowledge and perception of zoonotic diseases among hunters and wildlife traders in Epe, Lagos State, Nigeria. The increases in most of the variables evaluated and improvement in the overall knowledge base post-intervention serve as a pointer that our model (CAN-based intervention) could be adopted for behavioural change communication for zoonotic diseases, especially in resource-limited settings. Leveraging CAN strategies, individuals and communities can be empowered to make informed, health-related choices, thereby mitigating the risks associated with zoonotic diseases. Our findings of the role of CAN-based intervention in improving knowledge are in agreement with the previous work [25] on using integrated knowledge translation to move knowledge into action for more effective practice, programme and policy as a way to increase the relevance, applicability and impact of research.

Our model embraced the Information, Education, and Communication (IEC) approach, which has been acknowledged as a potent tool for catalysing essential shifts in understanding, attitudes, and behaviours [26]. The present study, through the video documentary and case studies, provided information, education and communication on zoonoses to the wildlife community in a participatory, interactive manner. As previously reported, videos are a type of IEC material that can be used to convey clear public health messages without relying on written or verbal explanations and to promote behavioural change [27]. Corrales et al. [28], in their study carried out in Medellin, Colombia, highlighted how a video intervention improved high school pupils’ awareness of zoonotic diseases spread by dogs. Again, the National Center for Emerging and Zoonotic Infectious Diseases (NCEZID) used animation and video to spread the word about its efforts to protect people from infectious disease dangers that exist both domestically and internationally [29]. These multi-media materials act as spotlights of knowledge, showcasing subjects like antibiotic-resistant diseases and the crucial One Health idea.

A systematic review [30] also supported the effectiveness of video-based interventions in changing knowledge and attitudes about infectious diseases. The video documentary as well as case studies employed in the present study among the wildlife hunters and traders not only raised awareness but could also provide important insights for policymakers useful in developing informed, all-inclusive mitigation strategies against zoonotic diseases.

Moreover, this study shows that, at pre-intervention, the majority did not believe and were not aware that humans could contract any diseases from wildlife. This observation is worrisome considering such an occupationally exposed group, coupled with the fact that up to 60% of emerging zoonotic diseases spread from domestic or wild animals to humans [2]. This incorrect position may spread to other people within the country, ultimately contributing to poor disease control [31]. This finding is similar to previous reports. For instance, Kusumaningrum et al. [32] reported a low level of awareness of zoonotic diseases among communities involved in the wildlife trade value chain in Indonesia.

Similarly, a study [33] showed that the majority of bushmeat hunters and traders studied indicated that diseases cannot be transmitted from bushmeat to humans in Nigeria. In contrast, however, a relatively higher level (55%) of zoonotic disease awareness was reported among Nigerian hunting communities [34]. Differences across study sites may be due to educational campaigns in the respective areas. We are unaware of public health outreach campaigns related to wildlife and disease in this present region of study.

Regarding some of the behaviours that could increase the risk of exposure of wildlife hunters and traders to zoonotic diseases, our findings reveal that most of the participants had experienced the situation of animals that died of unknown causes during hunting expeditions. The majority would take such animals home and process them for eating, as long as the carcass is still fresh. This kind of practice opens up the hunters and wildlife traders, as well as the unsuspecting public, including members of their families, to zoonoses. The risk of spillover may be associated with wild meat consumption [35], which is a global occurrence and is not just limited to wildlife markets and hunting practices in low-income and middle-income countries. Similarly, Wikramanayake et al. [36] developed a risk-assessment tool to evaluate disease risk presented by the food processing and handling of traded species in specific markets, therefore enabling the identification of high- risk trading conditions. Enhanced surveillance of human population groups, who could be more readily exposed through the consumption of at-risk animals and of the animal populations to which they are exposed, could help to predict spillover risk and enable early intervention [37]. Our finding is similar to a report [38] which indicated the practice of consumption of dead animals. According to these workers, carcass disposal among the Ilchamus was by consumption as it was against the culture to bury a dead animal. Often, this practice is reinforced by the notion that boiled meat is safe for consumption and could carry no disease [39].

Again, on the issues related to health and safety measures taken by the wildlife hunters and traders, all the participants had at least once or twice contact with wildlife animals. Despite being such a high-risk occupational group, the majority had not received any training on zoonotic diseases and their prevention. Worse still, about half of the participants had only occasionally or never gone for medical check-ups or health screenings. Self-care and home remedies have been associated with cultural beliefs and practices [40]. This is substantiated by the use of herbs and charms as part of the precautions taken by the hunters and wildlife traders in this study when handling or trading wildlife. Such cultural practices could result in a delay in treatment-seeking. Such cultural practices could also affect awareness and recognition of the severity of illness, availability of service and acceptability of service. It is very plausible that the lack of training on zoonotic diseases by the majority of these participants also contributes to the current practices among this occupational group. Judging by the significant differences observed following the interventions administered, it suffices to say that training this group of hunters and wildlife traders will go a long way in improving their understanding of zoonoses as well as positively redirecting their beliefs and practices.

Although the sample size was small, this was representative of the cohort in that setting. We believe the information obtained could be generalised to the larger population, given the cultural ties common to this set of people. In addition, the post-survey was immediately after the intervention. Future research would provide an opportunity to ascertain whether the result obtained will be sustained. The information obtained from the study will be useful to inform policy and the design of targeted interventions in the future.

## Conclusion

This study employed a Community Action Network-based intervention using a video documentary and case studies as model tools towards influencing the knowledge of zoonoses among wildlife hunters and traders in Epe, Lagos State, southwest Nigeria. The intervention significantly improved the knowledge of zoonoses among the studied population. Besides, the study documented important gaps in knowledge and cultural practices that could serve as a guide for the formulation of policies aimed at achieving effective control of zoonotic diseases in Nigeria. Considering the burden of zoonoses, particularly at the human-animal-wildlife interface, we propose the CAN-based intervention using a video documentary and case studies as a mitigation strategy towards raising the knowledge level and promoting behavioural changes regarding zoonotic diseases, especially among occupationally exposed communities in Nigeria and other sub-Saharan African countries.

## Data Availability

All relevant data are within the manuscript and its Supporting Information files

## Acknowledgments

We sincerely appreciate the support of the participants particularly the hunters who made this study possible. We acknowledge the support of the West Africa One-Health Consortium supported by the International Development Research Centre (IDRC) Canada.

## Author Contributions

C.S. conceived and designed the study. C.S., C.E., A.E.J., A.H.K., M,O., O,F., and F.E.O., developed the study survey tool. C.S., C.E., M.O., A.E.J., A.H.K., A.V.O., O.F., and F.E.O., carried out field work and data gathering. C,S., C.E., O.F., M.O., A.E.J., A.H.K., A.V.O., and A.K.O., carried out data analyses and visulization. A.H.K., A.E.J., C.E. conducted the quality appraisal and wrote the initial draft of the manuscript. C.S., O.D.O., R.A., and A.V.O., carried out a critical review of the manuscript and all authors approved the final version.

